# Diagnostic Accuracy of Global Longitudinal Strain and Speckle-Tracking-Derived Ejection Fraction for the Detection of Reduced Left Ventricular Ejection Fraction on Cardiac Magnetic Resonance: A Secondary Diagnostic Accuracy Analysis

**DOI:** 10.64898/2026.07.11.26357808

**Authors:** Álvaro Herrera-Escandón, Sebastián Ayala-Zapata, Stephany Barbosa-Balaguera

## Abstract

**Background:** Global longitudinal strain (GLS) and speckle-tracking-derived left ventricular ejection fraction (LVEF) are increasingly used to assess systolic function, but no consensus cut-off exists for identifying reduced LVEF as defined by cardiac magnetic resonance (CMR), the imaging reference standard. Our group previously reported excellent concordance between echocardiographic and CMR-derived LVEF in a cohort of 35 patients, but concordance statistics do not translate directly into clinically actionable diagnostic thresholds.

**Objectives:** To determine optimal cut-off values of GLS and speckle-tracking-derived LVEF for detecting CMR-defined reduced LVEF (<50%), and to compare their diagnostic accuracy, in a secondary analysis of this previously described cohort.

**Methods:** In this cross-sectional diagnostic accuracy study, 34 of the 35 originally enrolled patients (with complete strain and CMR data) were analyzed. Receiver operating characteristic (ROC) curve analysis estimated the area under the curve (AUC) for each index test against CMR-derived LVEF <50%. Optimal cut-offs were selected using the Youden index. Sensitivity, specificity, predictive values, and likelihood ratios were reported with 95% confidence intervals (CI). AUCs were compared using the DeLong test.

**Results:** Reduced LVEF (<50%) by CMR was present in 12 of 34 patients (35.3%). GLS showed an AUC of 0.892 (95% CI 0.729-0.996), with an optimal cut-off of -17.0% (sensitivity 75.0%, specificity 86.4%; positive likelihood ratio 5.50, negative likelihood ratio 0.29). Speckle-tracking-derived LVEF showed an AUC of 0.771 (95% CI 0.538-0.971), with an optimal cut-off of 52.8% (sensitivity 58.3%, specificity 95.5%). The difference in AUC was not statistically significant (DeLong p=0.192).

**Conclusions:** A GLS cut-off of -17.0% showed good discriminative accuracy for detecting CMR-defined reduced LVEF, numerically superior to but not statistically different from speckle-tracking-derived LVEF. These exploratory, hypothesis-generating findings require external validation in larger, multicenter cohorts before clinical application.

## Introduction

Left ventricular ejection fraction (LVEF) remains the principal parameter used to characterize systolic function and to guide diagnostic and therapeutic decisions in cardiovascular disease. Cardiac magnetic resonance (CMR) is considered the reference standard for LVEF quantification because of its high accuracy and reproducibility, but its cost and limited availability restrict its use as a first-line or serial test in most clinical settings, particularly in middle-income countries.

Global longitudinal strain (GLS), obtained by two-dimensional speckle-tracking echocardiography (2D-STE), has emerged as a complementary and more sensitive marker of myocardial deformation than LVEF, with demonstrated value in the detection of subclinical dysfunction and in prognostication across heart failure, cardiomyopathies, valvular disease, and cardio-oncology.

Our group previously reported concordance between Simpson-derived LVEF, GLS, and CMR-derived LVEF in a cohort of 35 patients evaluated at a cardiovascular clinic in Cali, Colombia, showing excellent agreement between Simpson-derived LVEF and CMR (Lin’s concordance correlation coefficient, CCC 0.831) and good agreement between GLS and CMR-derived LVEF (CCC 0.751) [1]. However, concordance coefficients such as the CCC quantify agreement between continuous measurements and are not designed to answer a distinct and clinically important question: given a single GLS or strain-derived LVEF value, how accurately can a clinician classify a patient as having reduced LVEF by CMR, and at what threshold? Answering this question requires a diagnostic-accuracy framework (cut-off, sensitivity, specificity, likelihood ratios, and predictive values) rather than a concordance analysis, and motivated the present secondary analysis of the same cohort.

Despite the increasing clinical use of GLS, no universally accepted cut-off exists for identifying reduced LVEF, and reported thresholds vary substantially depending on the clinical question being addressed. For example, a GLS cut-off of -20.0% has been proposed to identify patients with normal LVEF at risk of adverse outcomes [7], whereas a cut-off as low as -9.0% has been reported as optimal for predicting short-term clinical progression in stable heart failure [8]. This heterogeneity likely reflects differences in study populations, target conditions, reference standards, and vendor-specific strain software [9], and underscores the need for cut-off values derived specifically against CMR-defined LVEF categories in well-characterized cohorts.

Speckle-tracking-derived automated volumetric LVEF, obtained during the same acquisition as GLS, represents an additional, less studied index test that could theoretically track reference-standard LVEF more directly, but its comparative diagnostic performance against GLS for this purpose has not been well established.

The present study aimed to determine the diagnostic accuracy of GLS and speckle-tracking-derived LVEF for the detection of reduced LVEF (<50%) defined by CMR, to identify optimal cut-off values for each index test, and to compare their areas under the ROC curve, in a secondary analysis of a previously described cohort.

## Methods

### Study design and population

This was a cross-sectional diagnostic accuracy study, reported in accordance with the Standards for Reporting of Diagnostic Accuracy Studies (STARD 2015) where applicable to a secondary, exploratory analysis [10] (S1 Checklist). The population, recruitment procedures, and imaging acquisition protocols have been described in detail elsewhere [1]. In brief, patients aged 18 years or older with confirmed or suspected heart disease who underwent both a comprehensive transthoracic echocardiogram, including 2D-STE strain analysis, and CMR within a 24-hour interval at a cardiovascular clinic in Cali, Colombia, were considered eligible. Of the 35 patients originally enrolled, one was excluded from the present analysis because of incomplete strain and CMR volumetric data, leaving 34 patients. This is a secondary analysis of that cohort, addressing a diagnostic-accuracy question that was not examined in the original concordance-focused report.

### Index tests

Echocardiographic images were acquired in three standard apical views (two-, three-, and four-chamber) and analyzed using a dedicated speckle-tracking platform (TomTec). GLS was calculated as the average peak systolic longitudinal strain across the 17-segment left ventricular model. LVEF was independently derived from automated endocardial border detection during the same acquisition (speckle-tracking-derived LVEF). The echocardiographic operator was blinded to CMR results.

### Reference standard

CMR-derived LVEF, obtained from volumetric analysis of end-diastolic and end-systolic left ventricular volumes, served as the reference standard. CMR examinations were interpreted independently of the echocardiographic and strain results. Feature-tracking CMR strain analysis was not available during the study period and was not incorporated. For the primary analysis, CMR-derived LVEF was dichotomized at the 50% threshold, consistent with contemporary heart failure classification systems that distinguish preserved from reduced or mildly reduced ejection fraction [2].

### Statistical analysis

Continuous variables were tested for normality using the Shapiro-Wilk test and reported as mean ± standard deviation or median (interquartile range, IQR) as appropriate. Categorical variables were reported as absolute and relative frequencies, with the valid denominator specified whenever data were missing.

Diagnostic accuracy of GLS and speckle-tracking-derived LVEF for detecting CMR-defined LVEF <50% was assessed using ROC curve analysis. The AUC was calculated for each index test, with 95% CI obtained by non-parametric bootstrap resampling (5,000 replicates). Optimal cut-off values were selected using the Youden index (J = sensitivity + specificity - 1) [11]. At each optimal cut-off, sensitivity, specificity, positive predictive value (PPV), negative predictive value (NPV), and positive and negative likelihood ratios (LR+, LR-) were calculated; 95% CI for proportions used the Wilson score method [14] and 95% CI for likelihood ratios used the log method. The AUCs of GLS and speckle-tracking-derived LVEF were compared using the DeLong test for two correlated ROC curves [12], conceptually related to the earlier method of Hanley and McNeil [13].

A secondary threshold of CMR-derived LVEF <40% was also explored. After computing ROC estimates for this threshold, only 4 of 34 patients met this criterion, yielding an apparent AUC of 1.000; given that this reflects fewer than the commonly cited minimum of approximately 10 events per analysis for stable classification performance [15], we judged it too unstable for reliable interpretation and excluded it from the primary results. We report this as a post-hoc decision, made after observing the instability of the estimate rather than specified in advance, in the interest of full methodological transparency.

Because the optimal cut-off for each index test was derived and evaluated using the same sample, which can optimistically bias reported sensitivity and specificity [17], two internal validation procedures were performed as sensitivity analyses: (i) leave-one-out cross-validation (LOOCV), and (ii) Efron’s bootstrap optimism correction (2,000 resamples) [17]. A risk-of-bias and applicability assessment was additionally performed using the QUADAS-2 framework [18].

A two-sided p-value <0.05 was considered statistically significant. Analyses were performed in Python 3.12 using scikit-learn, statsmodels, and scipy.

## Results

### Study population

Of 35 originally enrolled patients, 1 (2.9%) was excluded because of incomplete strain and CMR volumetric data, leaving 34 patients (97.1%) for the analytic cohort (Fig 1). Reduced LVEF (<50%) by CMR was present in 12 of 34 patients (35.3%). No adverse events related to either the index tests or the reference standard occurred, as both are non-invasive imaging modalities.

**Fig 1.**
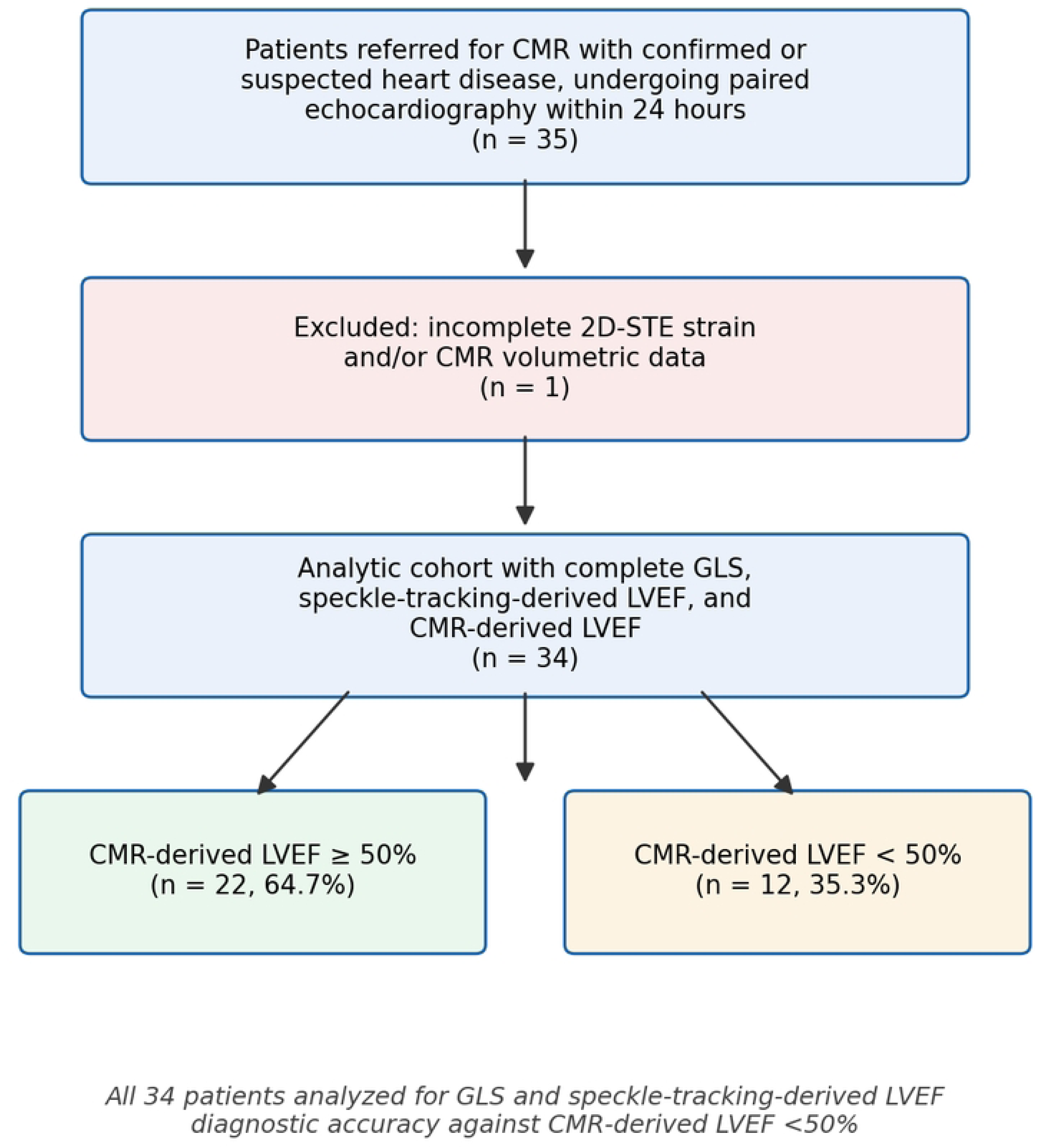
Patient flow diagram. CMR: cardiac magnetic resonance; GLS: global longitudinal strain; LVEF: left ventricular ejection fraction.

Baseline demographic and clinical characteristics of the analytic cohort are summarized in Table 1, and clinical indications for CMR in Table 2.

**Table 1.** Baseline demographic and clinical characteristics (n=34). IQR: interquartile range. *Denominator (N) reflects the number of patients with available data for that variable; comorbidity data were incomplete for a subset of patients (missingness 15-21% across variables). Laboratory biomarkers (troponin, NT-proBNP) were available in only 7/34 (20.6%) and 4/34 (11.8%) patients, respectively, and were therefore considered too incomplete for meaningful summary.

| Characteristic | Value |
| --- | --- |
| Age, years, median (IQR) | 56.5 (40.8-62.8) |
| Male sex, n (%) | 21 (61.8) |
| Hypertension, n/N (%)* | 14/27 (51.9) |
| Dyslipidemia, n/N (%)* | 10/28 (35.7) |
| Obesity, n/N (%)* | 9/28 (32.1) |
| Coronary artery disease, n/N (%)* | 6/28 (21.4) |
| Type 2 diabetes, n/N (%)* | 2/27 (7.4) |
| Smoking history, n/N (%)* | 3/29 (10.3) |

**Table 2.** Clinical indications for cardiac magnetic resonance (CMR) referral (n=34). CAD: coronary artery disease; MINOCA: myocardial infarction with non-obstructive coronary arteries.

| Clinical indication for CMR | n (%) |
| --- | --- |
| Suspected dilated cardiomyopathy | 9 (26.5) |
| MINOCA | 7 (20.6) |
| Suspected CAD / myocardial viability | 6 (17.6) |
| Hypertrophic cardiomyopathy | 5 (14.7) |
| Myocarditis | 4 (11.8) |
| Pericarditis | 1 (2.9) |
| Cardiac amyloidosis | 1 (2.9) |
| Not specified | 1 (2.9) |

**Table 3.** Descriptive statistics of GLS, speckle-tracking-derived LVEF, and CMR-derived LVEF (n=34). SD: standard deviation; IQR: interquartile range.

| Variable | Mean ± SD | Median (IQR) | Range |
| --- | --- | --- | --- |
| GLS, % | -17.5 ± 4.1 | -17.9 (-20.7 to -15.0) | -23.2 to -6.8 |
| Speckle-tracking-derived LVEF, % | 55.1 ± 13.7 | 58.8 (53.3-64.3) | 21.1-73.6 |
| CMR-derived LVEF, % | 53.5 ± 14.2 | 57.5 (44.2-64.0) | 20.5-75.0 |

**Table 4.** Area under the ROC curve (AUC) with 95% CI (bootstrap, 5,000 replicates) for GLS and speckle-tracking-derived LVEF for detecting CMR-derived LVEF <50%. DeLong test comparing the two AUCs: difference 0.121, Z=1.30, p=0.192.

| Index test | AUC (95% CI) | Optimal cut-off |
| --- | --- | --- |
| GLS | 0.892 (0.729-0.996) | > -17.0% |
| Speckle-tracking-derived LVEF | 0.771 (0.538-0.971) | < 52.8% |

**Table 5.** Diagnostic accuracy metrics (%) at the Youden-optimal cut-off for each index test, with 95% CI. Proportions use the Wilson score method; likelihood ratios use the log method. PPV: positive predictive value; NPV: negative predictive value; LR+: positive likelihood ratio; LR-: negative likelihood ratio.

| Index test (cut-off) | Sensitivity (95% CI) | Specificity (95% CI) | PPV (95% CI) | NPV (95% CI) | LR+ (95% CI) | LR- (95% CI) |
| --- | --- | --- | --- | --- | --- | --- |
| GLS (> -17.0%) | 75.0 (46.8-91.1) | 86.4 (66.7-95.3) | 75.0 (46.8-91.1) | 86.4 (66.7-95.3) | 5.50 (1.83-16.5) | 0.29 (0.11-0.78) |
| Strain-LVEF (< 52.8%) | 58.3 (32.0-80.7) | 95.5 (78.2-99.2) | 87.5 (52.9-97.8) | 80.8 (62.1-91.5) | 12.83 (1.78-92.4) | 0.44 (0.22-0.86) |

### Descriptive echocardiographic and CMR findings

GLS correlated significantly with CMR-derived LVEF (Pearson r=-0.72, p<0.001), as did speckle-tracking-derived LVEF (Pearson r=0.76, p<0.001).

### Diagnostic accuracy for detecting CMR-derived LVEF <50%

Cross-tabulation against the reference standard yielded, for the GLS threshold, 9 true positives, 3 false positives, 3 false negatives, and 19 true negatives. For speckle-tracking-derived LVEF, the classification was 7 true positives, 1 false positive, 5 false negatives, and 21 true negatives; the very wide interval around its positive likelihood ratio reflects the single false-positive classification. ROC curves for both index tests are shown in Fig 2.

**Fig 2.**
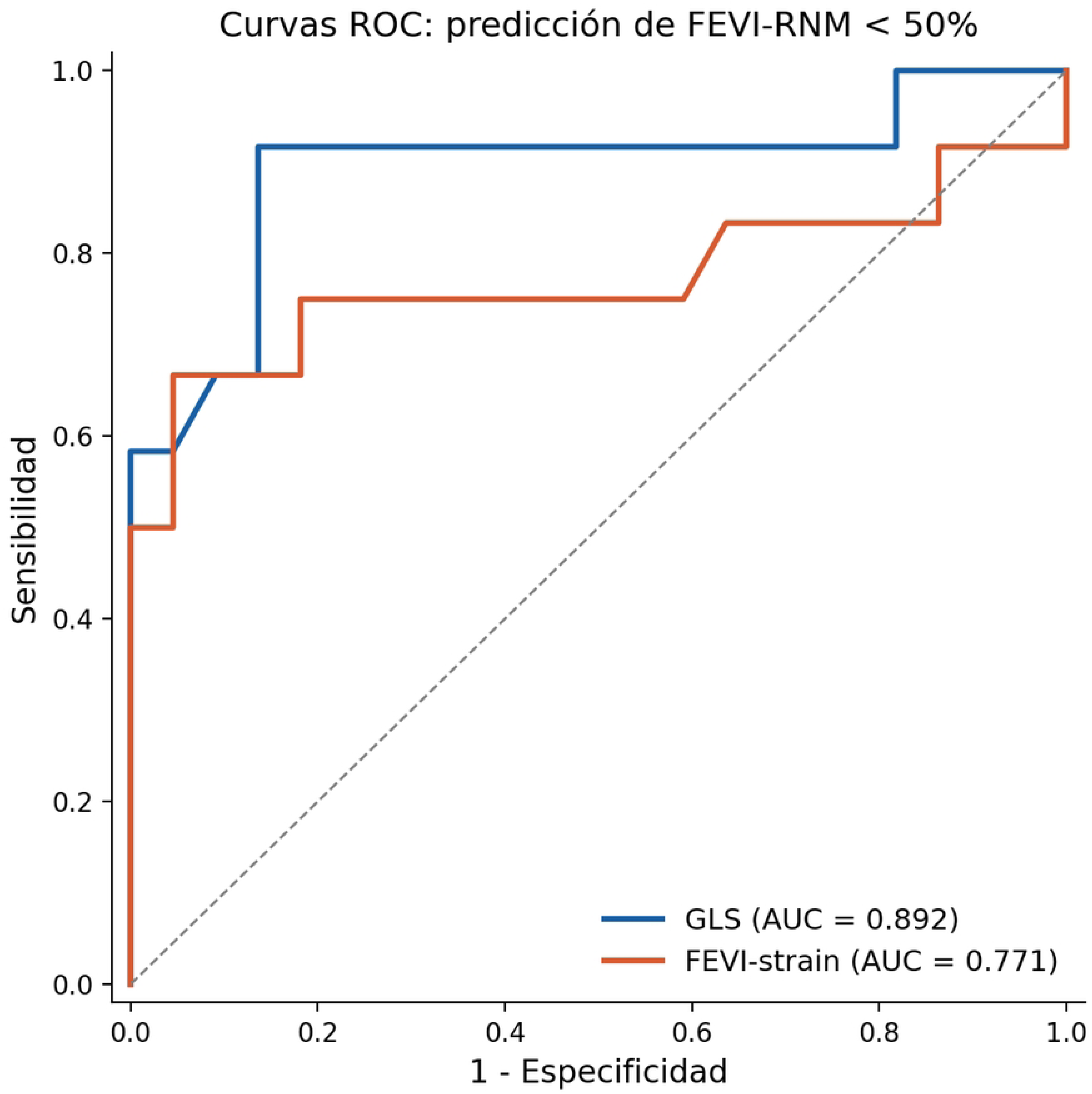
Receiver operating characteristic (ROC) curves for global longitudinal strain (GLS) and speckle-tracking-derived left ventricular ejection fraction (LVEF) for the detection of cardiac magnetic resonance-derived LVEF <50%.

### Internal validation of cut-off performance

Because the reported cut-offs were derived and evaluated on the same 34 patients, we assessed the magnitude of optimism using LOOCV and Efron’s bootstrap optimism correction [17] (Table 6). For GLS, the Youden-optimal cut-off was identical across all 34 leave-one-out iterations (-17.0%), and cross-validated sensitivity and specificity were unchanged from the apparent estimates. Bootstrap optimism correction likewise showed a negligible difference (<1 percentage point) between apparent and optimism-corrected estimates for both index tests. These findings suggest that, within the limits of this sample, the identified cut-offs are relatively stable and not primarily an artifact of overfitting; they do not, however, substitute for external validation in an independent cohort.

**Table 6.** Internal validation of cut-off performance. LOOCV: leave-one-out cross-validation; bootstrap-corrected estimates subtract the mean optimism across 2,000 bootstrap resamples (Efron’s method).

| Index test (cut-off) | Apparent sens. / spec. | LOOCV sens. / spec. | Bootstrap-corrected sens. / spec. |
| --- | --- | --- | --- |
| GLS (> -17.0%) | 75.0% / 86.4% | 75.0% / 86.4% | 75.3% / 85.7% |
| Strain-LVEF (< 52.8%) | 58.3% / 95.5% | 58.3% / 95.5% | 58.9% / 93.7% |

### Risk of bias and applicability (QUADAS-2)

A structured risk-of-bias assessment was performed using the QUADAS-2 framework [18] (Table 7). The main identified risk of bias was in the index-test domain: cut-offs for GLS and speckle-tracking-derived LVEF were derived empirically from the study data (Youden index) rather than pre-specified from prior literature, which can inflate apparent diagnostic accuracy. This risk is partially, but not fully, mitigated by the internal validation reported above. Risk of bias in the patient-selection, reference-standard, and flow-and-timing domains was judged low. Applicability concerns were rated as moderate given the heterogeneous, referral-based population.

**Table 7.** QUADAS-2 risk-of-bias and applicability assessment.

| Domain | Risk of bias | Applicability concern | Rationale |
| --- | --- | --- | --- |
| Patient selection | Low | Moderate | Consecutive eligible patients; heterogeneous referral population may limit generalizability to lower-prevalence settings |
| Index test | High | Low | Cut-off derived empirically (not pre-specified); partially mitigated by LOOCV/bootstrap internal validation |
| Reference standard | Low | Low | CMR is the accepted reference standard; interpreted independently of index test |
| Flow and timing | Low | - | Both tests within 24 hours; 34/35 (97%) included in analysis |

Fig 3 summarizes the key findings of the study.

**Fig 3.**
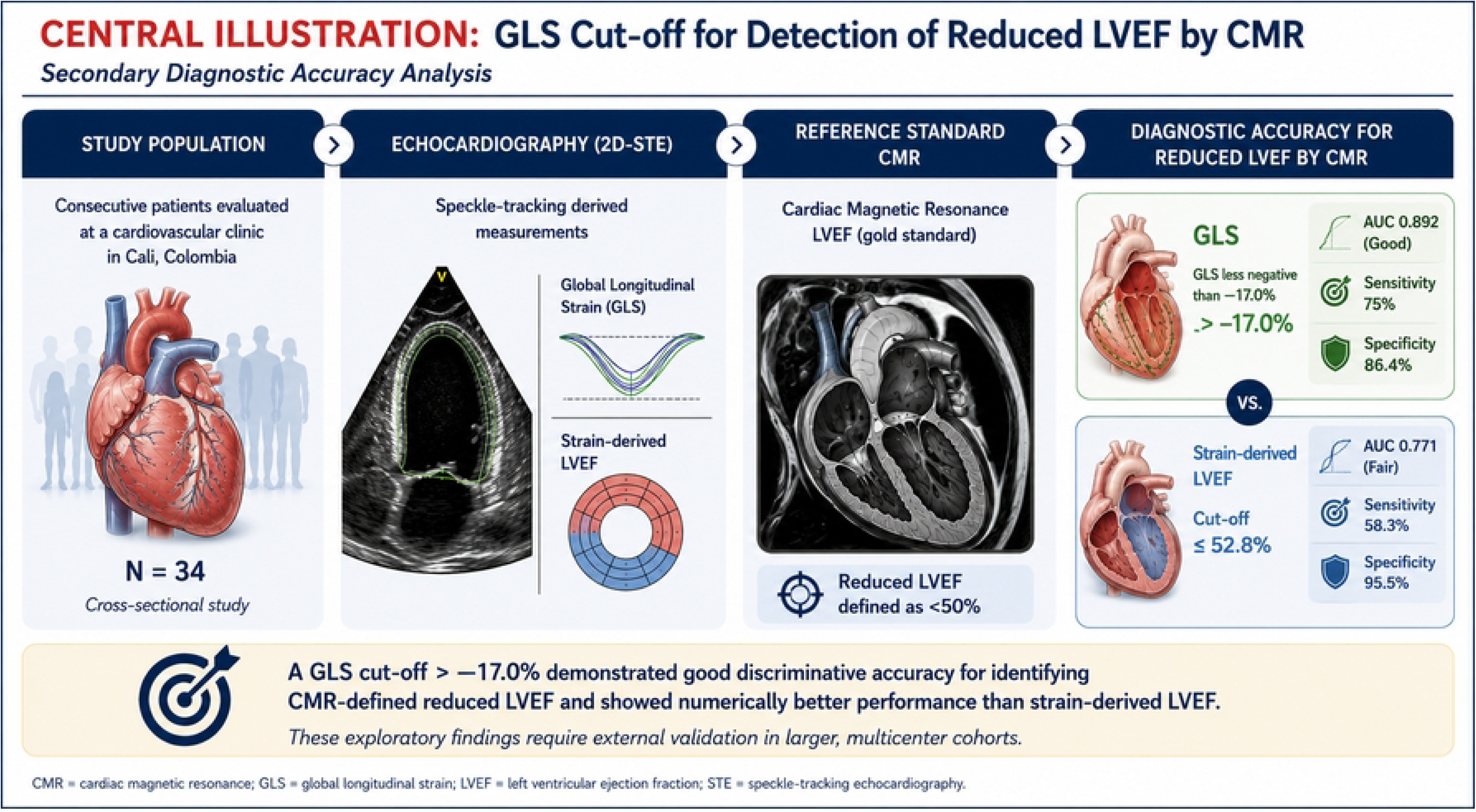
Central illustration. Graphical summary of the study: study population, echocardiographic index tests (GLS and speckle-tracking-derived LVEF), the CMR reference standard, and the diagnostic accuracy of each index test for detecting CMR-defined reduced LVEF (<50%). CMR: cardiac magnetic resonance; GLS: global longitudinal strain; LVEF: left ventricular ejection fraction; STE: speckle-tracking echocardiography.

## Discussion

In this secondary analysis of a well-characterized cohort undergoing both echocardiography and CMR, a simple echocardiographic strain threshold identified patients with CMR-defined reduced LVEF. A GLS value less negative than -17.0% detected reduced LVEF (<50%) with good-to-excellent accuracy (AUC 0.892) and performed at least as well as automated speckle-tracking-derived LVEF (AUC 0.771). In a clinical setting where CMR is often unavailable, this suggests that a single, routinely obtainable strain value can help flag patients whose systolic function is likely to be reduced on the reference standard.

The relative advantage of GLS over speckle-tracking-derived LVEF has a plausible physiological basis. Longitudinal subendocardial fibers, which GLS interrogates directly, are typically the first to be affected by ischemic, infiltrative, or toxic myocardial injury, often before global chamber-level volumetric changes become apparent. Speckle-tracking-derived LVEF, by contrast, still depends on endocardial border tracking and geometric assumptions similar to those underlying Simpson’s method, which may explain both its closer behavior to conventional volumetric indices and its comparatively lower discrimination for a CMR-defined binary outcome in this cohort.

Our optimal GLS threshold is consistent with the range reported in the wider literature. Benyounes et al. found that two-dimensional GLS accurately detected reduced LVEF in a large series of unselected patients undergoing routine echocardiography, supporting the feasibility of a strain-based threshold for identifying impaired systolic function [19]. At the same time, published GLS cut-offs vary widely with the clinical question, from around -20.0% proposed to identify patients with normal LVEF at risk of adverse outcomes [7] to values as low as -9.0% for predicting short-term progression in stable heart failure [8]. This heterogeneity reflects differences in populations, target conditions, reference standards, and vendor-specific strain software [9], and reinforces the value of thresholds derived specifically against CMR-defined LVEF categories, as attempted here. These results complement, rather than contradict, our group’s earlier concordance analysis of the same cohort, in which GLS showed good but not excellent agreement with CMR-derived LVEF as a continuous variable [1]. Concordance and discrimination answer different questions: a marker can have only moderate absolute agreement with a reference standard, limiting its use for interchangeable reporting of an LVEF value, while still ranking patients accurately enough to support a binary clinical decision. In practical terms, a test that should not be used to report an interchangeable LVEF number may still be well suited to flagging patients who likely have reduced LVEF.

From a health-systems perspective, particularly relevant in Latin American settings where CMR access is limited by cost and availability, a validated GLS threshold could serve as a pragmatic triage tool: patients with GLS less negative than -17.0% could be prioritized for confirmatory CMR or closer follow-up, while those with more preserved strain could reasonably be managed with echocardiographic surveillance. This role is supported by the likelihood ratios observed: a positive GLS result raised the odds of CMR-defined reduced LVEF roughly five-fold, while a negative result substantially lowered them. Importantly, with a sensitivity of 75% at this threshold, approximately one in four patients with true reduced LVEF would be missed by GLS alone, so the threshold should complement rather than replace clinical judgment and direct volumetric assessment when reduced LVEF is otherwise suspected.

An important caveat for clinical translation is that GLS values and thresholds vary meaningfully across vendors and software platforms; identical recordings analyzed with different systems can yield systematically different absolute strain values [9]. Because GLS in this study was measured on a single platform, the -17.0% threshold should not be assumed to transfer directly to other vendors without local recalibration or harmonization. Two statistical considerations temper these findings. The numerically higher accuracy of GLS over speckle-tracking-derived LVEF did not reach statistical significance, and this comparison was underpowered; a future confirmatory study would require on the order of 155 to 210 patients to detect a difference of this magnitude [13,16]. In addition, because the thresholds were derived and evaluated in the same small sample, internal validation was used to gauge their stability and suggested minimal optimism, but this does not replace external validation in an independent cohort.

## Limitations

This study has several limitations. First, the sample size (n=34, a fixed cohort with no possibility of further recruitment) is small, yielding wide confidence intervals and limited power; the cut-off values should be considered exploratory and hypothesis-generating. Second, the cut-offs were derived empirically from this dataset rather than pre-specified, a recognized source of bias; although internal validation suggested minimal optimism, this cannot substitute for external validation. Third, a secondary threshold of CMR-derived LVEF <40% was explored but not reported as a primary result because only 4 patients met this criterion. Fourth, the single-center design, the use of a single strain platform, and the clinical heterogeneity of the population may limit generalizability, given known inter-vendor variability in strain measurements. Fifth, comorbidity data were missing for 15-21% of patients across several variables, and laboratory biomarkers were available in only a small minority. Finally, feature-tracking CMR strain was not available for comparison, and the cross-sectional design does not allow assessment of the prognostic value of the proposed cut-off.

## Conclusions

In this exploratory secondary analysis, a GLS cut-off of -17.0% showed good-to-excellent accuracy for detecting CMR-defined reduced LVEF (<50%), numerically higher than but not statistically different from a speckle-tracking-derived LVEF cut-off of 52.8%. These findings should be externally validated in larger, multicenter cohorts, ideally with pre-specified sample sizes powered for diagnostic accuracy comparisons, before the proposed cut-off is adopted in clinical practice.

## Ethics statement

This study was a secondary analysis of a retrospectively assembled cohort. It was approved by the Ethics Committee of the Universidad del Valle and Hospital Universitario del Valle, Cali, Colombia (approval code 058-2024, December 2024). Given the retrospective nature of the analysis and the use of de-identified data, the Ethics Committee granted a waiver of the requirement for informed consent.

## Data availability

All relevant data are within the paper and its Supporting Information files.

## Funding

The authors received no specific funding for this work.

## Competing interests

The authors have declared that no competing interests exist.

## Author contributions

Conceptualization: Álvaro Herrera-Escandón, Stephany Barbosa-Balaguera. Data curation: Álvaro Herrera-Escandón. Formal analysis: Sebastián Ayala-Zapata. Methodology: Sebastián Ayala-Zapata. Investigation: Stephany Barbosa-Balaguera. Writing, original draft: Stephany Barbosa-Balaguera. Writing, review and editing: Álvaro Herrera-Escandón, Sebastián Ayala-Zapata, Stephany Barbosa-Balaguera. Project administration: Stephany Barbosa-Balaguera.

## Relationship to prior publication

This manuscript reports a secondary, diagnostic-accuracy analysis of a cohort previously described by our group in a concordance study among Simpson-derived LVEF, GLS, and CMR-derived LVEF [1]. The present analysis addresses a distinct question (optimal cut-off, sensitivity, specificity, likelihood ratios, and predictive values for a dichotomized reference standard) that was not examined in that report. This overlap has been disclosed to the editor in the cover letter, in accordance with ICMJE recommendations on overlapping publications.

## Use of Artificial Intelligence

During the preparation of this work, the authors used a generative artificial intelligence tool (ChatGPT, OpenAI; image-generation feature, 2026) to create the central illustration (Fig 3), a schematic summary of the study design and main findings. No research data, results, statistical analyses, or manuscript text were generated by artificial intelligence. The authors reviewed the illustration against the study data for accuracy, edited it as needed, and take full responsibility for its content.

## Supporting information

**S1 Checklist.** STARD 2015 reporting checklist.

## Data Availability

All relevant data are within the paper and its Supporting Information files.

## Notes

### Competing Interest Statement

The authors have declared no competing interest.

### Author Declarations

Ethics Committee of the Universidad del Valle and Hospital Universitario del Valle, Cali, Colombia. Approval number 058-2024 (December 2024). The committee granted a waiver of informed consent given the retrospective nature of the analysis and the use of de-identified data.

